# Episode-Driven Insights: Can Large Language Models Tackle Multimodal Diabetes Data?

**DOI:** 10.1101/2025.04.24.25326385

**Authors:** Hee Jung Choi, Shriti Raj

## Abstract

This study explores the potential of state-of-the-art large language models (LLM) to scaffold type 1 diabetes management by automating the analysis of multimodal diabetes device data, including blood glucose, carbohydrate, and insulin. By conducting a series of empirically grounded data analysis tasks, such as detecting glycemic episodes, clustering similar episodes into patterns, identifying counterfactual days, and performing visual data analysis, we assess whether models like ChatGPT 4o, Claude 3.5 Sonnet, and Gemini Advanced can offer meaningful insights from diabetes data. Our findings show that ChatGPT 4o demonstrates strong potential in accurately interpreting data in the context of specific glycemic episodes, identifying glycemic patterns, and analyzing patterns. However, limitations in handling edge cases and visual reasoning tasks highlight areas for future development. Using LLMs to automate data analysis tasks and generate narrative summaries could scaffold clinical decision-making in diabetes management, which could make frequent data review feasible for improved patient outcomes.

## Introduction

Type 1 diabetes (T1D) is a complex condition associated with severe health complications, hospitalizations, and premature mortality. Its lifelong management requires continuous and precise monitoring of complex data from multiple sources, including continuous glucose monitors (CGM), insulin pumps, and dietary records^1^. Despite the availability of these rich data streams, interpreting them to inform disease management remains a significant challenge for both patients and healthcare providers^2^. Frequent engagement with wearable data, although can improve glycemic outcomes, is impractical because of the high effort associated with analysis and interpretation of data^3^. Current digital platforms distill multidimensional device data in complex visualizations that impose a significant cognitive and temporal burden on patients and clinicians, limiting effective review of data in short clinic visits, resulting in clinician burnout and reduced patient engagement^3^. Ineffective engagement with data despite available data platforms results in missed opportunities for care and suboptimal blood glucose control^4^. With increasing uptake of CGMs and insulin pumps, clinicians and patients must frequently navigate complex datasets to make timely decisions. Informatics tools that improve the interpretation and accessibility of data can empower them both to enhance diabetes management.

Recent work by Raj et al. highlighted the promise of episode-driven narratives in facilitating data comprehension for T1D care^5^. Their research demonstrated how narratives that contextualize glycemic episodes with potential causal factors can significantly reduce the cognitive load on both patients and clinicians. However, generating these narratives manually is labor-intensive and requires expert time, suggesting the need to explore automated approaches that are scalable and that can model expert knowledge and thinking. This presents an opportunity to explore the use of large language models (LLMs) in automating the generation of episode-driven narratives. LLMs, which have demonstrated remarkable capabilities in processing and generating complex natural language content in diverse contexts, could offer a novel solution for synthesizing multimodal diabetes data into relatively more intuitive, and comprehensible formats, such as natural language narratives^6, 8^. For instance, data visualizations when accompanied by natural language narratives are better understood by readers as compared to the visualizations alone^8^. Furthermore, a recent work by Healey et al. demonstrated the feasibility of using LLMs to generate accurate and safe summaries of CGM data^7^. While this work established the potential of LLMs in handling medical time series data, it focused on a single data stream, i.e., blood glucose levels. Considering multimodal data is particularly critical in T1D management, as it allows for examining blood glucose levels by considering different influential factors, such as carbohydrate intake, and insulin administration. The integration of multimodal data into narratives can ease the burden that is associated with visual analysis of multiple data streams to identify trends and correlations. Work by Alavi et al. demonstrated the feasibility of using LLMs with multimodal wearable data on research tasks to make new discoveries^9^. We build on these prior works to investigate the potential of using state-of-the-art (SOTA) LLMs to interpret multimodal diabetes data in not only textual but also visual form.

We conduct a series of experiments using synthetic data generated by publicly available T1D data generator and SOTA LLM-powered chat interfaces to assess their ability to perform key tasks such as hyper- and hypo-glycemic episode detection, clustering, exploratory analysis, and graph interpretation. In particular, we used OpenAI’s ChatGPT 4o, Anthropic’s Claude 3.5 Sonnet, and Google’s Gemini Advanced (1.5 Pro) in our analyses^10, 11, 12^. We focus on five key tasks that reflect real-world clinical needs: episode detection (hypoglycemia and hyperglycemia identification on individual days), episode clustering to identify recurring patterns, counterfactual day detection, statistical analysis, and visual reasoning (interpreting CGM plots alongside carbohydrate and insulin data). Our objective is to establish whether current SOTA LLMs can handle these tasks effectively and explore their potential for future integration into informatics tools for diabetes data review.

## Methods

### Design

This study is a proof-of-concept aimed at exploring the ability of SOTA LLMS to interpret multimodal T1D data through a series of structured Chain of Thought (CoT) tasks. Simulated CGM, carbohydrate (CHO), and insulin data were generated using “simglucose,” a publicly available Python implementation of the FDA-approved UVa/Padova Simulator (2008 version)^13^. We did so due to the privacy concerns associated with feeding real patient data into public LLMs, as was also done by Healey et al^7^. We generated 7-day CGM, CHO, and insulin data for 10 adult patients, which were then used in various prompt-driven tasks. The use of simulated data is crucial for protecting patient privacy while still providing a valid foundation for evaluating LLM performance in interpreting multimodal diabetes data.

The study involved evaluating the performance of SOTA LLMs across five distinct tasks designed to simulate real-world T1D data interpretation scenarios^14^. Each task was structured to assess specific capabilities of the models, including their ability to detect glycemic episodes, cluster episodes by time windows, detect counterfactual days, perform statistical analyses on the detected episodes, and perform visual reasoning using charts. These tasks are fundamental to generating clinically relevant narratives building on the concept of episode-driven narratives as described by Raj et al^5^. To ensure that the analysis remains as updated as possible, these models—ChatGPT 4o, Claude 3.5 Sonnet, and Gemini Advanced (1.5 Pro)—were evaluated on September 15-16, 2024. Outputs were assessed based on correctness, as consistency is critical to automating the interpretation process.

#### Task A: Episode Detection Task

The first task required the LLMs to identify periods of suboptimal glucose management by detecting continuous episodes of hypoglycemia (glucose < 70 mg/dL) and hyperglycemia (glucose > 180 mg/dL). The LLMs were prompted to analyze the start and end times of these episodes, ensuring proper identification of breaks when glucose levels returned to the target range (70-180 mg/dL). This step is foundational for subsequent tasks that build on the detection of episodes to identify patterns by compiling recurring occurrences.

The evaluation for Task A was based on a point system that assessed the accuracy of detecting hyperglycemia and hypoglycemia episodes. LLMs were awarded 1 point for correctly identifying the total number of hyperglycemia episodes and 1 point for correctly identifying the total number of hypoglycemia episodes. Additionally, 1 point was given for *each* hyperglycemia episode where *both* the start and end times were accurate, and another point was awarded for *each* hypoglycemia episode with correct start *and* end times. The overall accuracy score for Task A was calculated by dividing the total number of correctly identified elements by the maximum possible points. If the answer was not presented in the correct format, 0 points were assigned for that particular component.

#### Task B: Episode Clustering Task

Once episodes were detected, the LLMs were tasked with clustering them into recurring occurrences based on predefined time windows (e.g., early morning, midday, late evening, etc.). This is critical in identifying problematic glycemic trends over several days. Identifying these repeated occurrences allows for focused treatment decisions.

The evaluation for Task B was based on identifying clusters of episodes, where a cluster was defined by both a predefined time window and the type of episode (hyperglycemia or hypoglycemia). LLMs were awarded 1 point for each correctly identified cluster, with the requirement that both the time window and the episode type were accurately matched. The accuracy score for Task B was calculated by dividing the total number of correctly identified clusters by the total possible clusters. If any component of the cluster (time window or episode type) was incorrect, 0 points were assigned for that cluster.

#### Task C: Counterfactual Days Detection Task

In this task, the LLMs were asked to identify “counterfactual days”—days when the opposite condition of a pattern occurred. For instance, if midday hypoglycemia was detected as a recurring pattern, the LLMs were prompted to identify days where there was no hypoglycemia during the midday window. This helps identify features that differentiate the two types of days, hinting at possible changes that could be made^15^.

The evaluation for Task C was based on the LLMs’ ability to correctly identify counterfactual days. For *each* cluster identified in Task B, 1 point was awarded if all counterfactual days were accurately identified. To earn a point, the model needed to correctly list the “Did not occur on” days for each cluster as specified in the prompt (see Task C of Figure 1). The accuracy score for Task C was calculated by dividing the number of correctly identified clusters by the total number of clusters. If any counterfactual days were incorrect or missing, 0 points were assigned for that cluster.

**Figure 1.**
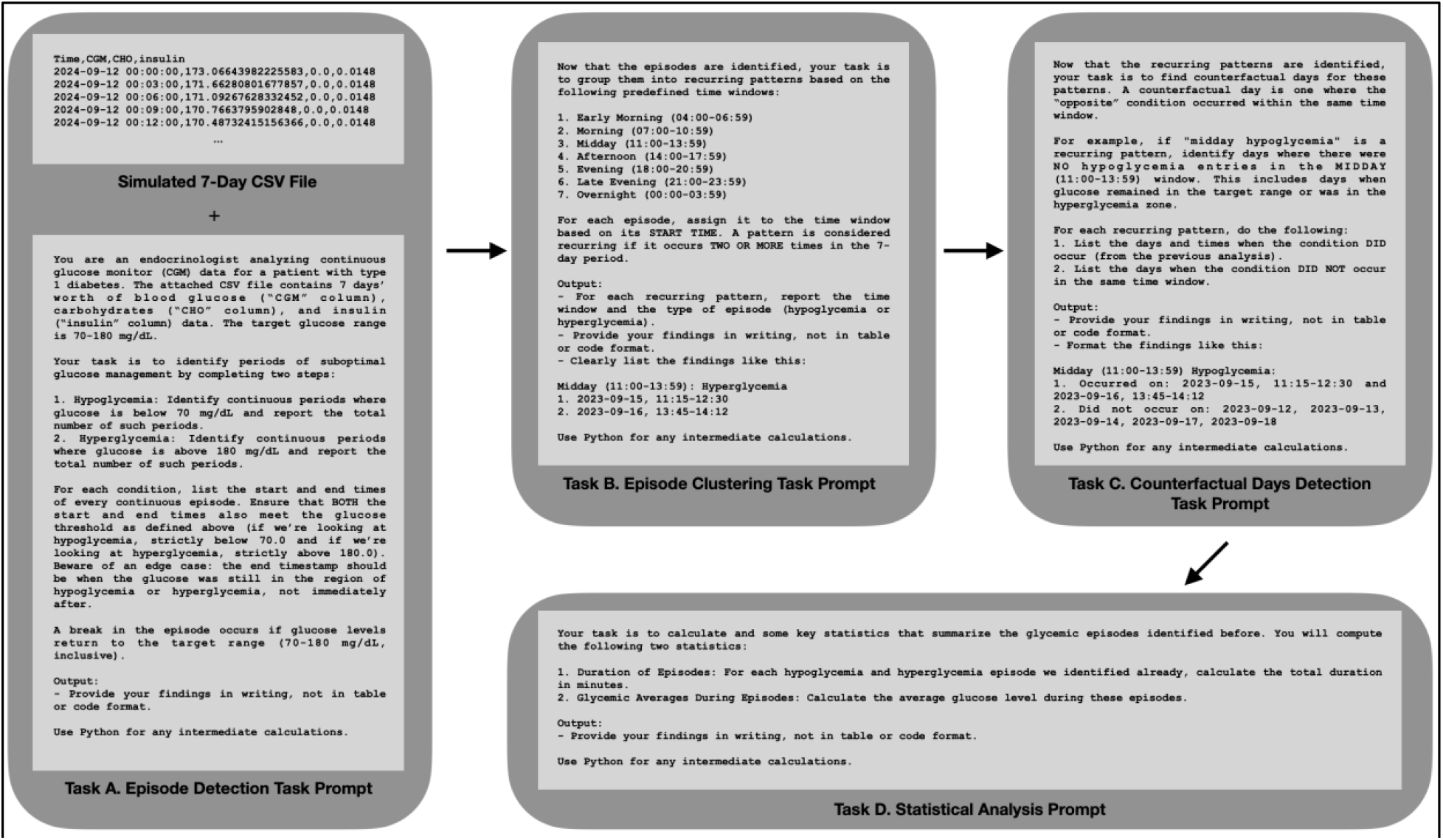
The CoT prompting processed is illustrated above. We initially feed in a 7-day T1D CSV file and prompt step-by-step.

#### Task D: Statistical Analysis Task

In Task D, the focus was on the statistical analysis of the identified hypoglycemia and hyperglycemia episodes, specifically calculating the duration of each episode and the average glucose level during these periods. This extends the work of Healey et al., who demonstrated the feasibility of using LLMs to compute broader glycemic metrics such as glucose variability, Time in Range (TIR), and the Glucose Management Indicator (GMI). Instead of replicating these existing measures, we chose to focus on episode-specific metrics—duration and average glucose—because they provide critical insights into the severity and frequency of glycemic excursions^7^. Understanding the duration of extreme glucose episodes allows for prioritization based on the potential impact on patient outcomes, while the average glucose during each episode offers a more granular understanding of how far the glucose deviated from the target range^16^.

The evaluation for Task D was based on the LLMs’ ability to perform statistical analysis on the identified episodes of hypoglycemia and hyperglycemia. Points were awarded as follows: 1 point for each episode where the total duration was correctly calculated, and 1 point for each episode where the average glucose level was accurately calculated. The total score was the sum of points for all episodes. The accuracy score for Task D was calculated by dividing the total number of correctly calculated elements by the maximum possible points. If any calculation was incorrect, 0 points were assigned for that episode.

The prompts for Tasks A-D are illustrated in Figure 1.

#### Task E1: Visual Reasoning Task

Given that visual interpretation of data displays is an integral part of T1D care as it happens today, Task E1 evaluated the LLMs’ ability to perform visual reasoning. Five randomly selected 1-day datasets (glucose, carbohydrate, and insulin data) from different patients were presented as visual plots, as seen in Figure 2. The LLMs were prompted to identify episodes of hypoglycemia and hyperglycemia, and relevant carbohydrate intake, and bolus insulin administration details based on the attached plots.

**Figure 2.**
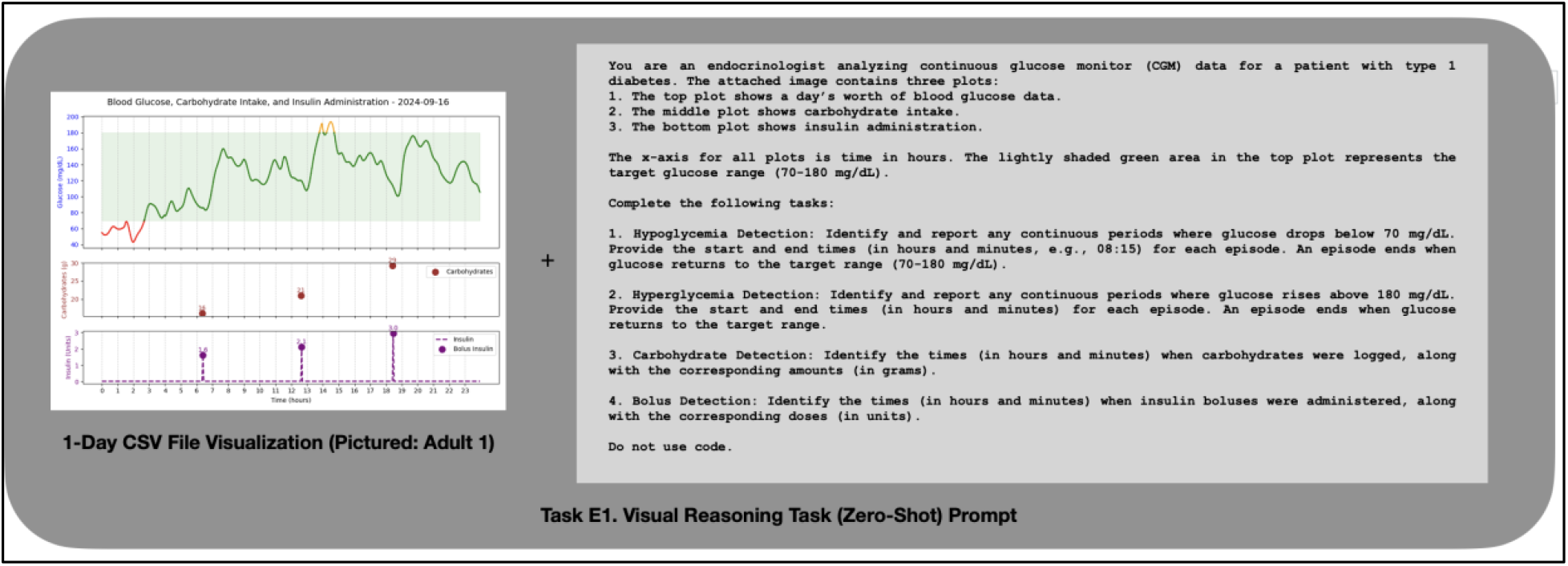
Task E1 prompting is done by feeding in both image and text.

#### Task E2: Visual Reasoning Task with One-Shot Prompting

In Task E2, the LLMs were presented with a one-shot example where step-by-step reasoning was demonstrated before asking them to analyze new, unseen plots. The goal was to assess whether LLMs could generalize the visual reasoning approach demonstrated in Task E1 and apply it to fresh data. The decision to explore one-shot prompting exclusively in Task E2, rather than in the other tasks, was based on the understanding that prior research has already established the strong performance of LLMs in zero-shot and few-shot contexts for textual data^17^. However, visual data, particularly in the context of diabetes, has been less explored in these prompting strategies. Given the complexity of interpreting multimodal visual information like CGM plots, we aimed to investigate whether a one-shot approach, with step-by-step reasoning, could enhance the model’s ability to generalize its visual reasoning to new, unseen data.

For both Tasks E1 and E2, the evaluation was based on the LLMs’ ability to accurately identify hyperglycemia and hypoglycemia episodes, carbohydrate intake, and insulin bolus entries based on visual input. For each hyperglycemia and hypoglycemia episode, 1 point was awarded if both the start and end times were accurately identified within 30 minutes of the ground truth. This margin of leniency was introduced to account for the inherent difficulty in pinpointing exact times from graphical data. If no episodes were present, 1 point was given for correctly stating that there were none. For carbohydrate intake, 1 point was assigned for each correctly identified entry, requiring both the carbohydrate amount and the time to match the ground truth, with the time being within 30 minutes of accuracy. Similarly, for insulin bolus entries, 1 point was awarded for each correct identification, with the bolus dose and time needing to be within 30 minutes of the ground truth. The total accuracy score for Tasks E1 and E2 was calculated by dividing the sum of correctly identified elements by the total possible points across all categories.

### Main Outcome Measurement

The main outcome of the study was the correctness of the LLMs’ outputs in completing each task. Correctness was evaluated by comparing the models’ outputs against the expected results (ground truth) derived from the simulated data. Each task was designed to produce standardized, interpretable outputs, ensuring consistency and ease of comparison across the three LLM platforms.

Ground truth data was generated using a custom script that automatically calculated the necessary values for comparison. To ensure the accuracy of the script, its output was cross-verified by the first author through triangulation with visualizations of the simulated T1D data. An example of this visualization is available in Figure 2 and Figure 3. Additionally, clustering and statistical analysis were manually performed and calculated to further confirm the correctness of the script and the derived ground truth. This triangulation ensured the reliability of the ground truth used for evaluation.

**Figure 3.**
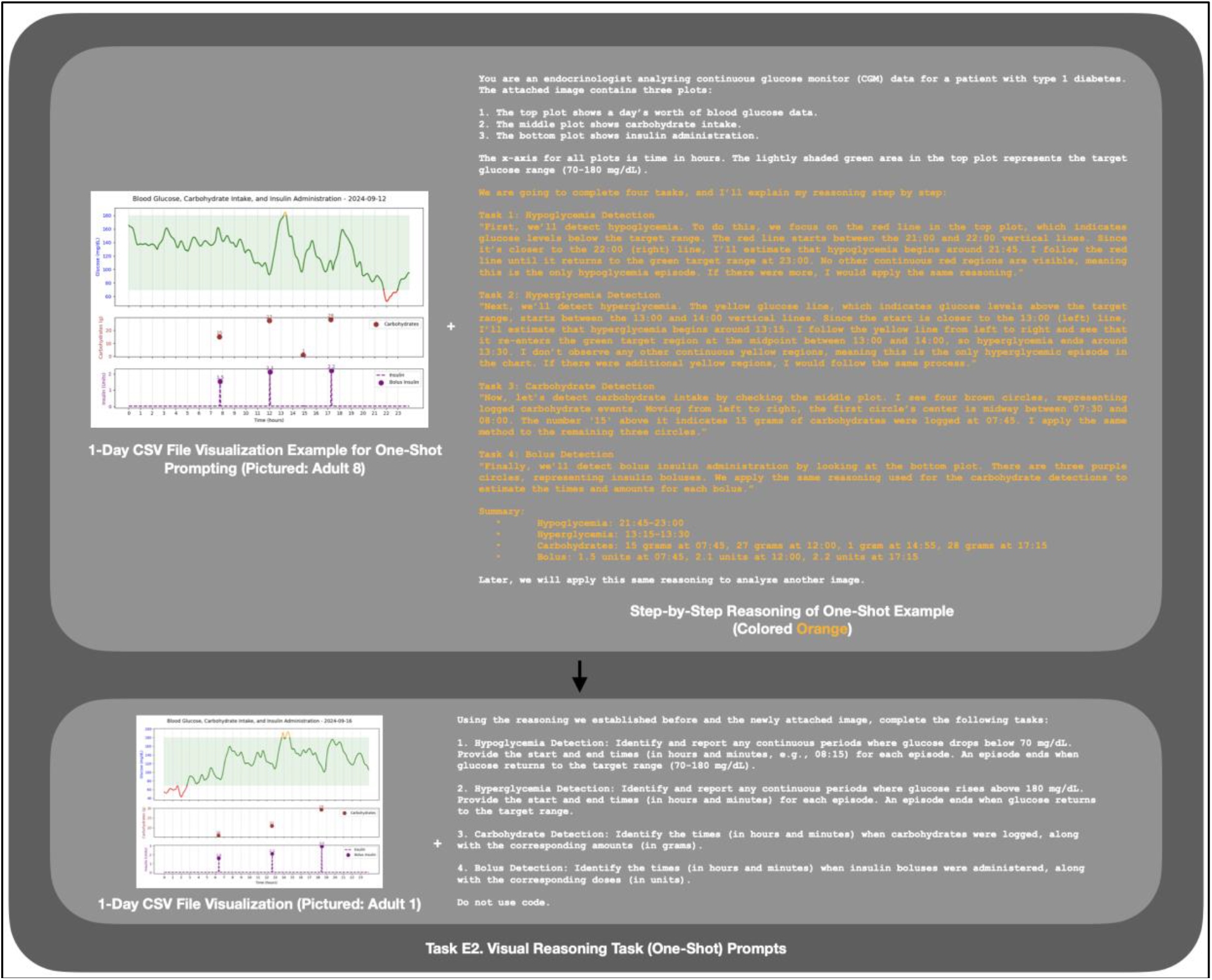
Task E2 is conducted by first providing the LLMs with a step-by-step reasoning example, then assessing their ability to generalize this approach to analyze new, unseen CGM, carbohydrate, and insulin data plots.

## Results

For certain patients, some tasks were marked as “N/A” due to the stepwise evaluation process used in this study. Task A (Episode Detection) served as the foundational task, and only if the LLM correctly identified the episodes in Task A did it proceed to the subsequent tasks. If the model failed Task A, no further evaluation was conducted, and the results for Tasks B, C, and D were automatically marked as “N/A.” Similarly, for Task B (Episode Clustering), if the clusters were not correctly identified, the model did not proceed to Task C (Counterfactual Days Detection) and was instead evaluated directly on Task D (Statistical Analysis). This stepwise approach is essential because all tasks were designed as CoT prompts. Each task built on the correctness of the previous one, meaning that errors in earlier stages would render subsequent analyses less meaningful.

### Task A: Episode Detection

In Task A, ChatGPT 4o outperformed the other models with a mean accuracy of 0.78 across 10 patients. However, it displayed variability, with near-perfect accuracy for most patients but a sharp decline in accuracy for some. The sharp decline in ChatGPT 4o’s accuracy for some patients was due to the model generating code that did not correctly handle the edge case where the end timestamp should be within the hypoglycemic or hyperglycemic range, resulting in time windows that were offset by certain amounts. Gemini Advanced (1.5 Pro) achieved relatively high performance for several patients. Claude 3.5 Sonnet performed very poorly overall, frequently hallucinating incorrect episodes. This meant that it could not proceed to the remaining tasks.

**Table 1.** Accuracy by model for task A.

| Patient | 1 | 2 | 3 | 4 | 5 | 6 | 7 | 8 | 9 | 10 | Mean |
| --- | --- | --- | --- | --- | --- | --- | --- | --- | --- | --- | --- |
| **ChatGPT 4o** |  |  |  |  |  |  |  |  |  |  |  |
| Accuracy | 1.0 | 1.0 | 1.0 | 1.0 | 1.0 | 0.0870 | 0.5 | 0.182 | 1.0 | 1.0 | **0.777** |
| **Claude 3.5 Sonnet** |  |  |  |  |  |  |  |  |  |  |  |
| Accuracy | 0.0417 | 0.0 | 0.0 | 0.0 | 0.0 | 0.0435 | 0.0 | 0.0 | 0.0 | 0.0 | **0.00852** |
| **Gemini Advanced (1.5 Pro)** |  |  |  |  |  |  |  |  |  |  |  |
| Accuracy | 0.0417 | 0.167 | 1.0 | 0.957 | 0.0 | 0.0 | 1.0 | 1.0 | 1.0 | 0.0800 | **0.525** |

### Task B: Episode Clustering

ChatGPT 4o performed consistently well, achieving perfect accuracy for most patients. Many of the entries here are N/A, as Task A was done incorrectly for a patient, in which case we chose not to have the model proceed with the next task.

**Table 2.** Accuracy by model for Task B.

| Patient | 1 | 2 | 3 | 4 | 5 | 6 | 7 | 8 | 9 | 10 |
| --- | --- | --- | --- | --- | --- | --- | --- | --- | --- | --- |
| **ChatGPT 4o** |  |  |  |  |  |  |  |  |  |  |
| Accuracy | 1.0 | 1.0 | 1.0 | 1.0 | 1.0 | N/A | N/A | N/A | 0.8 | 1.0 |
| **Claude 3.5 Sonnet** |  |  |  |  |  |  |  |  |  |  |
| Accuracy | N/A | N/A | N/A | N/A | N/A | N/A | N/A | N/A | N/A | N/A |
| **Gemini Advanced (1.5 Pro)** |  |  |  |  |  |  |  |  |  |  |
| Accuracy | N/A | N/A | 0.5 | N/A | N/A | N/A | 1.0 | 1.0 | 0.8 | N/A |

### Task C: Counterfactual Days Detection

ChatGPT 4o once again showed strong performance. Accuracy was only evaluated for models that accurately completed Task B for a given patient, as it is crucial to complete Task B correctly for Task C to be accurate.

**Table 3.** Accuracy by model for Task C.

| Patient | 1 | 2 | 3 | 4 | 5 | 6 | 7 | 8 | 9 | 10 |
| --- | --- | --- | --- | --- | --- | --- | --- | --- | --- | --- |
| **ChatGPT 4o** |  |  |  |  |  |  |  |  |  |  |
| Accuracy | 1.0 | 1.0 | 1.0 | 0.0 | 1.0 | N/A | N/A | N/A | N/A | 1.0 |
| **Claude 3.5 Sonnet** |  |  |  |  |  |  |  |  |  |  |
| Accuracy | N/A | N/A | N/A | N/A | N/A | N/A | N/A | N/A | N/A | N/A |
| **Gemini Advanced (1.5 Pro)** |  |  |  |  |  |  |  |  |  |  |
| Accuracy | N/A | N/A | N/A | N/A | N/A | N/A | 1.0 | 0.0 | N/A | N/A |

### Task D: Statistical Analysis Task

Again, ChatGPT 4o maintained its strong performance, achieving near-perfect accuracy.

**Table 4.** Accuracy by model for Task D.

| Patient | 1 | 2 | 3 | 4 | 5 | 6 | 7 | 8 | 9 | 10 |
| --- | --- | --- | --- | --- | --- | --- | --- | --- | --- | --- |
| **ChatGPT 4o** |  |  |  |  |  |  |  |  |  |  |
| Accuracy | 1.0 | 1.0 | 1.0 | 0.0 | 1.0 | N/A | N/A | N/A | 1.0 | 1.0 |
| **Claude 3.5 Sonnet** |  |  |  |  |  |  |  |  |  |  |
| Accuracy | N/A | N/A | N/A | N/A | N/A | N/A | N/A | N/A | N/A | N/A |
| **Gemini Advanced (1.5 Pro)** |  |  |  |  |  |  |  |  |  |  |
| Accuracy | N/A | N/A | 0.5 | N/A | N/A | N/A | 1.0 | 0.5 | 0.5 | N/A |

### Task E1: Visual Reasoning (Zero-Shot)

Task E1 focused on evaluating the LLMs’ visual reasoning capabilities using a single-day CGM plot for five randomly selected patients. ChatGPT 4o achieved a mean accuracy of 0.519. The model performed better when data points were clearly distinguishable (like the CHO and insulin data, which were all conspicuously indicated by a large dot, along with a numerical label) but struggled with continuous glucose data. Claude 3.5 Sonnet had a similar mean accuracy but showed a slight drop in consistency across the other patients. Like ChatGPT, it performed well with discrete data points but encountered difficulties with continuous time-series data.

**Table 5.**
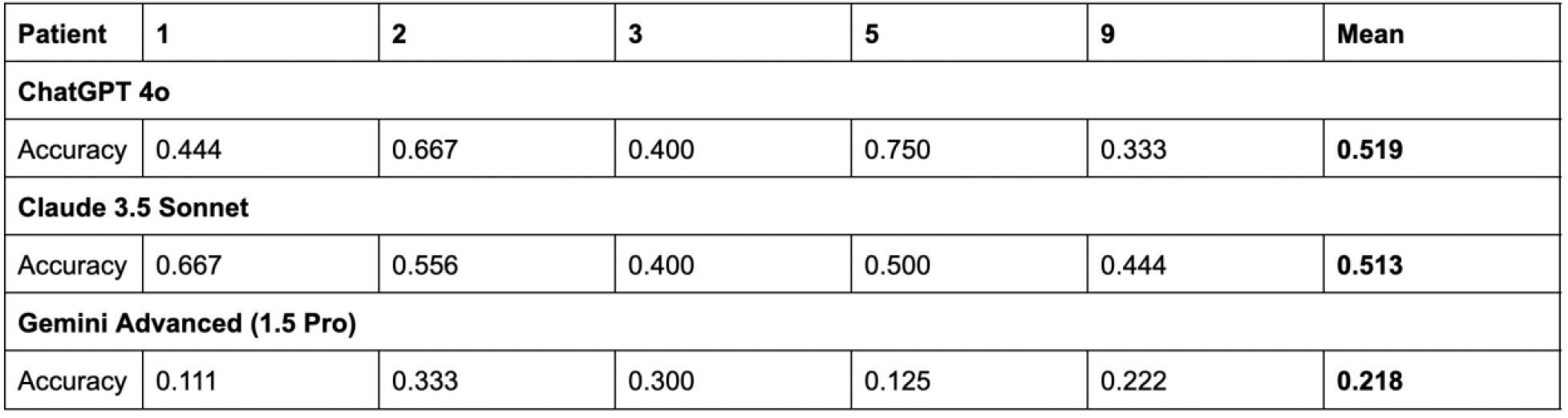
Accuracy by model for Task E1.

### Task E2: Visual Reasoning (One-Shot Prompting)

In Task E2, the same visual reasoning task was conducted with one-shot prompting. The overall improvement was marginal, as the model still encountered challenges with time-series interpretation. One-shot prompting provided marginal improvements for ChatGPT 4o but mostly had little effect across all models.

**Table 6.** Accuracy by model for Task E2.

|  | 1 | 2 | 3 | 5 | 9 | Mean |
| --- | --- | --- | --- | --- | --- | --- |
| **ChatGPT 4o** |  |  |  |  |  |  |
| **Accuracy** | 0.444 | 0.333 | 0.400 | 0.875 | 0.556 | **0.522** |
| **Claude 3.5 Sonnet** |  |  |  |  |  |  |
| **Accuracy** | 0.667 | 0.556 | 0.200 | 0.375 | 0.444 | **0.448** |
| **Gemini Advanced (1.5 Pro)** |  |  |  |  |  |  |
| **Accuracy** | 0.111 | 0.333 | 0.300 | 0.250 | 0.111 | **0.221** |

## Discussion

This study demonstrated the feasibility of using SOTA LLMs for interpreting multimodal T1D data. Among the tested models—OpenAI’s ChatGPT 4o, Anthropic’s Claude 3.5 Sonnet, and Google’s Gemini Advanced (1.5 Pro)— ChatGPT 4o consistently outperformed the others, particularly in episode detection and statistical analysis of the episodes. However, even the strongest model showed limitations in handling specific edge cases and visual reasoning with time-series data, highlighting important areas for improvement in future LLM applications.

The decline in accuracy for certain tasks, especially for ChatGPT 4o in Task A, was primarily due to its inability to handle edge cases related to timestamp detection in hypoglycemic or hyperglycemic episodes. Oftentimes, the model would output an end timestamp that was immediately after the hypoglycemic or hyperglycemic range, resulting in time windows that were offset by certain amounts. This limitation suggests that while LLMs show promise in automating T1D data interpretation, they still require more robust handling of continuous data and complex conditions either through human intervention or more precisely crafted prompts.

This study represents one of the first to comprehensively evaluate different SOTA LLMs across a range of tasks directly applicable to multimodal medical time series data for T1D care. The use of CoT prompting to standardize task outputs allowed for a structured comparison across models and offered insight into their current capabilities and limitations. The episodic analysis performed in Tasks A through D establishes a foundational framework for analyzing multimodal diabetes data using LLMs, providing a pathway for further biomedical applications, such as clinical decision support systems. Given the benefits and challenges of frequent diabetes data reviews, LLM-driven automation of data analysis and insight generation can lay the path for scalable solutions for frequent data-driven treatment intensification in the management of T1D.

In addition to demonstrating the feasibility of using LLMs to interpret multimodal medical time series data, a key contribution of our work is the incorporation of visual reasoning tasks (Task E), which are crucial for clinical applications in T1D care. Clinicians rely heavily on CGM plots to interpret patient data, and while models like ChatGPT 4o showed some capability in zero-shot and one-shot visual reasoning, the overall performance of all tested models was limited. This highlights the need for further refinement in LLMs’ ability to interpret time-series data visualizations, which are commonly used for medical data.

The ability of LLMs to assist in analyzing T1D data has direct implications for reducing clinician workload and enhancing patient care. As denoted by prior work, episode-driven narratives^5^ can simplify the interpretation of complex glucose, carbohydrate, and insulin data streams, leading to easier engagement with data. By automating parts of episode-based analysis, such as providing statistical insights on episodes, and representing these insights in natural language, LLMs could enable more efficient decision-making and can make these data more accessible to patients, who may not have the skills to interpret the data.

This study is a proof of concept, and several limitations must be addressed in future work. First, the simulated data used in this study may not fully represent the complexities of real-world clinical data. While the decision to use simulated data was necessary to avoid privacy concerns, future studies should validate the models on real patient data under controlled conditions. Another limitation is the inconsistent performance across tasks, particularly in visual reasoning and edge-case handling. Future work should focus on improving LLMs’ ability to interpret continuous time-series data more accurately, particularly in visual formats, which are essential for T1D care. Additionally, incorporating more advanced reasoning techniques and refining the models’ ability to handle edge cases will be crucial for improving their robustness.

## Conclusion

This study provides evidence on the feasibility of using LLMs to analyze multimodal diabetes device data from continuous glucose monitors and insulin pumps. Using an episode-driven approach, it shows that LLMs can accurately identify glycemic episodes and interpret other relevant data streams in the context of those episodes. An episode-driven approach to analysis and interpretation of diabetes data using LLMs can provide a framework for automating diabetes data analysis and insight generation.

## Data Availability

All data produced in the present study are available upon reasonable request to the authors.

